# Infant Weight Faltering and Feeding Patterns at 6 Weeks Post-Delivery in a Rural East African Community:Comparing Incremental Versus Static Weight Assessments

**DOI:** 10.1101/2023.12.13.23299669

**Authors:** Adenike Oluwakemi Ogah

**Affiliations:** University of Zambia

**Keywords:** incremental and static weight assessment, weight faltering, weight acceleration, exclusive breastfeeding, post-partum maternal depression score

## Abstract

**Background:** Early stages of infant weight faltering is difficult to detect in the first 6 months of life. Major reason for this might be the method of assessing infant growth. Incremental growth assessment, which is rarely performed in developing countries, is more useful than static growth assessment in infants. In a remote, understudied village in East Africa, this study examined the prevalence and determinants of infant weight faltering and compared incremental with static weight assessments among 6 weeks old infants.

**Subject and methods:** This was a secondary analysis of a prospective cohort data. Data of 512 out of the 529 mother-newborn pairs recruited from birth were obtained and analysed at 6 weeks post-delivery in the postnatal clinic. Records of infant feeding patterns and anthropometry were documented. Infant weight at birth and at 6 weeks were compared using the NICE criteria for both incremental/interval and static weight growth. Mothers were interviewed using the Edinburgh postpartum depression score to assess emotional status. Chi test and binary logistic regression model was used to examine the relationship between maternal/infant characteristics and infant weight growth. The results were presented in p-values, Odds ratio and 95% confidence interval. The similarity and dissimilarity between infant static and incremental weight assessments were measured using Kappa and McNemar tests respectively.

**Results:** Overall, the incidences of static and incremental weight faltering at 6 weeks post-delivery were 3.1% (16 out of 512) and 1.4% (7 out of 512), respectively. The Cohen Kappa measure of agreement between the 2 methods of weight assessment, was moderate at 0.424 (p<0.001). 3 out of the 512 infants were not exclusively breastfed; 2 of whom were offered water and 1 was fed with fresh cow milk.

A higher percentage of the SGA-born (compared to AGA and LGA) infants, 11 out of 107 (10.3%) were weight faltering according to static assessment. This was contrary to increment weight assessment, where a lower percentage of SGA-born [only 2 out 107 (1.9%), compared to LGA] infants, were faltering. The LGA-born infant, according to incremental weight assessment, was the least likely to weight accelerate, compared to the SGA-born and AGA-born infants; OR 0.04; 95% CI 0.01, 0.10. High maternal depression score was associated with infant weight acceleration, p<0.001; OR 1.13; 95% CI 1.06, 1.21. Boys were less likely to weight accelerate compared to girls, p<0.001; OR 0.45; 95% CI 0.29, 0.70. Infants, who were adequately fed were more likely to weight accelerate compared to those who were poorly fed, p<0.001; OR 2.80; 95% CI 1.55, 5.05. Infants, who did not fall ill since birth were more likely to weight accelerate compared to those who had fallen ill, p=0.004; OR 2.73; 95% CI 1.37, 5.43.

**Conclusion and Recommendations:** This study emphasizes the importance of assessing infant growth using both static and incremental measures. Health workers need to be trained to carry out incremental growth assessment in infants. Lactational and mental support programs should be strengthened in the rural MCH systems, to assist mothers to achieve pleasant experiences with breastfeeding and newborn care. Exclusive breastfeeding should be encouraged and this will in turn reduce the incidence of infant ill health. Home visits should be carried out for infants lost to follow up.

## Background

Growth faltering (previously known as failure to thrive) is a broad term used to describe children, who fall below their anticipated growth trajectory due to undernutrition. The term ‘growth faltering’ refers to static growth, or an unexpected downward trajectory and/or unexpected slower rate of weight gain from a previously established pattern of growth.^1^

According to the National Instituite for Health and Care Excellence (NICE) criteria, concerns about weight faltering should be entertained, when an infant’s current body weight is below the 2^nd^ centile for age, regardless of the birthweight.^1^ This definition was based on static (one-time) growth assessment, which suggests attained growth and does not include or it minimizes changes in growth trajectories over time.^2^ Because infant growth is dynamic and their growth pattern is exponiential rather than linear, especially in the first 6 months of life, incremental (interval) growth assessment is more appropriate rather than static assessment in this early age group. Relying only on static growth assessment (whether one-time or serially), as it is being implemented currently, in many developing countries, could lead to misdiagnosis or late diagnosis of growth faltering in this age group. The static assessment might remain the same over a period of time, while the incremental or interval growth assessment may reveal earlier, an accelerated, flat or decelerated growth trend.^2^

According to the principles of human growth, postnatal growth occurs in discontinuous saltatory spurts with a stagnant background in between those spurts. Also growth is highly variable and maximum during early infancy, compared to other phases of postnatal human growth.^3^ Small children tend to move upwards through the centiles, while large children tend to cross downwards, following the normal phenomenon of regression to the mean and conditional weight gain.^4^

Growth faltering is caused primarily by inadequate feeding. Only 5-10% of cases are caused by underlying disease and another 5% caused by neglect-related issues.^5^ Olusanya and Renner observed that the growth findings from the Centers for Disease Control and Prevention (CDC) reference and World Health Organization (WHO)’s multicenter growth standards (based on static one-time measures such as weight-for-length, weight-for-age and length-for-age z-scores), differred at birth and within the first 3 months of life among 445 full term singleton in Lagos West Africa. Weight faltering was considered if weight-for-age percentile was below 3. The authors reported the percentages of infants growth faltering to be 20.7% at birth and declined to 16.4% at follow-up, on any of the growth charts. LBW-born infants were significantly more likely to falter at followup.^6^ Boys were more prone to weight faltering than girls. Maleta et al. observed that length faltering was present at birth and persisted for the first 3 years of life, while weight faltering was at its peak between 3 and 12months of life among 767 live born babies in rural Malawi.^7^

The adverse effects of early postnatal growth faltering on the developing infant have been well documented and includes subsequent persistent growth faltering, increased risk of severe infection, delay in motor, cognitive and psychomotor development, behavioural problems, learning disabilities and infant mortality, to mention a few.^8^ Hence, it is crucial to use valid tools to regularly and accurately assess growth, identify early stages of growth faltering and intervene appropriately and effectively in this vulnerable population. The paucity of literature on infant incremental growth assessment is an indication that health professionals do not always carry out comprehensive growth assessment. Hence, this study was designed to determine the incidence and risk factors of growth faltering and to compare both increment and static weight assessments among 512 infants in a rural East African community.

## Concept of the study

Figure 1, shows the maternal and neonatal determinants of infant weight faltering and acceleration, that were investigated in this study. Static and incremental weight criteria were used to measure infant growth.

**Fig. 1:**
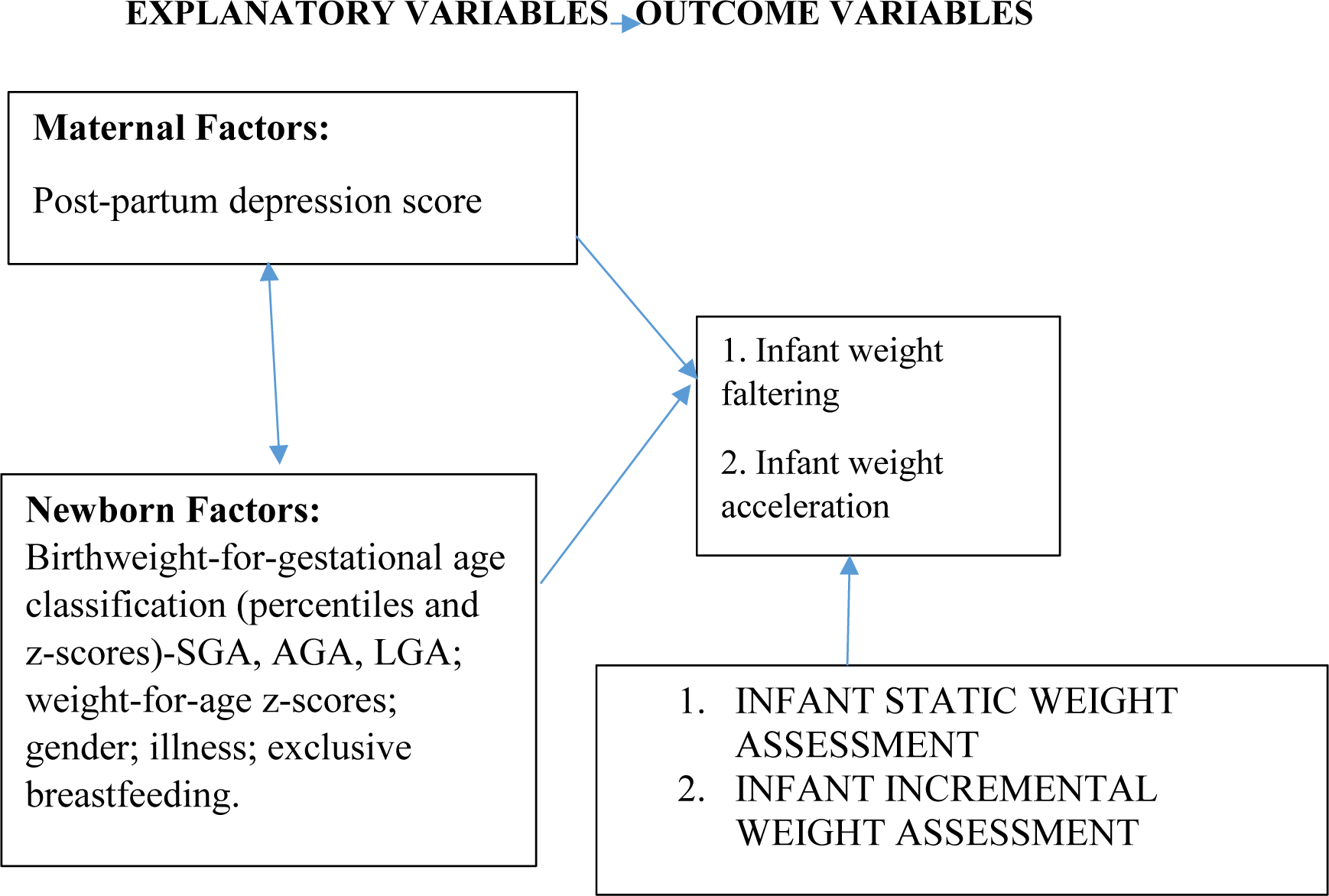
Concept of the study.

## Materials and methods

The methods employed in carrying out this study is discussed in this section.

### Study setting

Rwanda is a low-income, agricultural and landlocked country with approximately 11 million people living in five provinces, covering an area of 26,338 km^2^. It is called the home of a ‘1000 hills’^9^. The limited area of flat land available in most part of Rwanda is a hindrance to farming, animal rearing and construction of standard residential houses, among others. For example, the recommended minimum of 50 feet distance between source of household water and sewage tank in residential yards are often compromised during construction, leading to contamination of water source. There are 2 peak raining seasons in the country: April to June and September to November.

Rwanda has an average of 4.4 persons per household and a gross domestic product per capita of US $780.80. About half (48%) of its population is under 19 years of age and 39% live below the poverty line with a life expectancy at birth of 71.1 years for women and an adult literacy rate of 80% among 15–49 years old women. In addition, 87.3% of the population have health insurance and access to health services; spending an average of 47.4 min to reach a health centre.

Non-availability of regular supplies of clean and safe water, has been a longstanding problem in Rwanda, probably as a result of its landlocked and hilly terrains, making construction and supply of piped water, a major challenge. The public piped water flow infrequently and the taps and pipes may be rusted and breached in some places, especially in the rural areas, further contributing to contamination of household water. Many families store rain water in big tanks for use in their homes and this may become polluted (in the writer’s opinion) because of difficulties of cleaning these storage tanks. A few non-profit organizations, such as USAID and Water-for-life have sunk bore holes in strategic locations in a few number of villages in the country, with the aim of alleviating this water problem.

Gitwe village is located on a high altitude of 1,674 meters above sea level, in the southern province, 240km from Kigali, which is the capital city of Rwanda. Gitwe general hospital began in 1995, immediately after the genocide, for the purposes of providing medical services and later training to this isolated community. The hospital currently has 100% government support, since year 2020. The maximum number of deliveries at the hospital per month was about 200 and about 50 infants visit the maternal and child health clinic (MCH) per day. There are only 2 immunisation days per week.

Some of the challenges in the hospital include poor specialist coverage and few trained health workers, poor supply of equipment, water, electricity, laboratory services and medicines. Challenging cases are referred to the University of Rwanda Teaching Hospital in Butare or Kigali. Gitwe village was selected for this study, because there was no published birth data from this poorly researched, remote, relatively unaccessible community. In 2019, birth and up to 9 months of age feeding and growth data on 529 healthy mother-singleton newborn pairs were compiled in this village, over a period of 12 months in the delivery, postnatal wards and clinics of Gitwe general hospital and at its annex, the MCH.^9^

### Data source and sample

This was a secondary analysis of data collected for a prospective cohort study on growth faltering over a period of 12 months, until infant was 9 months old. Mother-newborn pairs were recruited consecutively, on first-come-first-serve basis. Maternal file review and newborn anthropometry [weight (kg), length (cm) and head circumference (cm) measurements, their z-scores and percentiles, were recorded to the nearest decimals] were carried out, soon after birth. Newborn gestational age was determined using the record of the maternal first date of the last menstrual period (LMP), fetal ultrasound dating and/or expanded new Ballard criteria. Newborn classification into small-for-gestational age (SGA), appropraite-for-gestational age (AGA) and large-for-gestational age (LGA) was based on their weight-for-gestational age percentiles at birth, according to WHO. Weight faltering at 6 weeks post-delivery was defined by NICE static weight assessment of weight-for-age z-score of ≤2. Weight faltering (according to NICE incremental weight faltering criteria), was considered when there was a fall across 1 or more weight centile spaces, if birthweight was below the 9th centile; when there was a fall across 2 or more weight centile spaces, if birthweight was between the 9th and 91st centiles; when there was a fall across 3 or more weight centile spaces, if birthweight was above the 91st centile; or when the current weight is ≤ 2nd centile for age, whatever the birthweight. One centile space on the percentile curve was equivalent to 0.67 z-scores.^1^

Weight acceleration by incremental weight assessment was documented, when the weight-for-age z-score at 6 weeks was more than 0.67 above that of the weight-for-gestational age z-score at birth. Weight-for-age z-score at birth and 6 weeks was obtained from PediTool calculator, which was based on WHO growth standards and used the infants’ corrected age.^10^

Mothers were interviewed using standardized infant feeding questionnaires and the Edinburgh postpartum depression questionnaire/score at 6 weeks post-partum in the postnatal clinic.^11^ The questionnaires were read to the mothers and filled by the research assistants.

To ensure the quality of data collected, 2 registered nurses were trained as research assistants at Gitwe Hospital for 2 days on the over-all procedure of mother and newborn anthropometry and data collection by the investigator. The questionnaires were pre-tested before the main data collection phase, on 10 mother-infant pair participants (2% of the total sample). The investigator closely followed the day-to-day data collection process and ensured completeness and consistency of the questionnaires administered each day, before data entry.

### Statistical analysis

Data clean up, cross-checking and coding were done before analysis. These data were entered into Microsoft Excel statistical software for storage and then exported to SPSS version-26 for further analysis. Both descriptive and analytical statistical procedures were utilized. Participants’ categorical characteristics were summarised in frequencies and percentages. Chi test was used to determine association between infant characteristics and weight faltering. Numerical characteristics such as maternal depression scores were presented in median and interquartile ranges for skewed data. A binary logistic regression model was created to examine the relationships between the independent variables (maternal and neonatal characteristics) and dependent variables (weight faltering and weight acceleration) and to generate the odds ratio. Factors with p-values <0.1 were included in the regression model. Odds ratio (OR), with a 95% confidence interval (CI) were computed to assess the strength of association between independent and dependent variables. For all, statistical significance was declared at p-value < 0.05. The reporting in this study were guided by the STROBE guidelines for observational studies.^12^

### Ethics

The Health Sciences Research Ethics Committee of the University of the Free State in South Africa gave the ethical approval for the collection of the primary data for the original study-’Growth and growth faltering in a birth cohort in rural Rwanda: a longitudinal study’ with Ethical Clearance Number: UFS-HSD2018/1493/2901. Written permission to collect data was obtained from the Director of Gitwe Hospital, Rwanda and the eligible mothers gave their informed consent before enrolment. The participants were given research identity numbers and the principal investigator was responsible for the safe keeping of the completed questionnaires and collected data, to ensure anonymity and confidentiality of the participants.

## Results

The following are the results obtained from the study.

### Participants

Five hundred and ninety-seven (597) babies were delivered at Gitwe Hospital, Rwanda, between 3^rd^ January and 9^th^ May 2019, out of which, eligible 529 mother-newborn pairs were enrolled into the prospective cohort study at birth, Figure 2.

**Fig 2:**
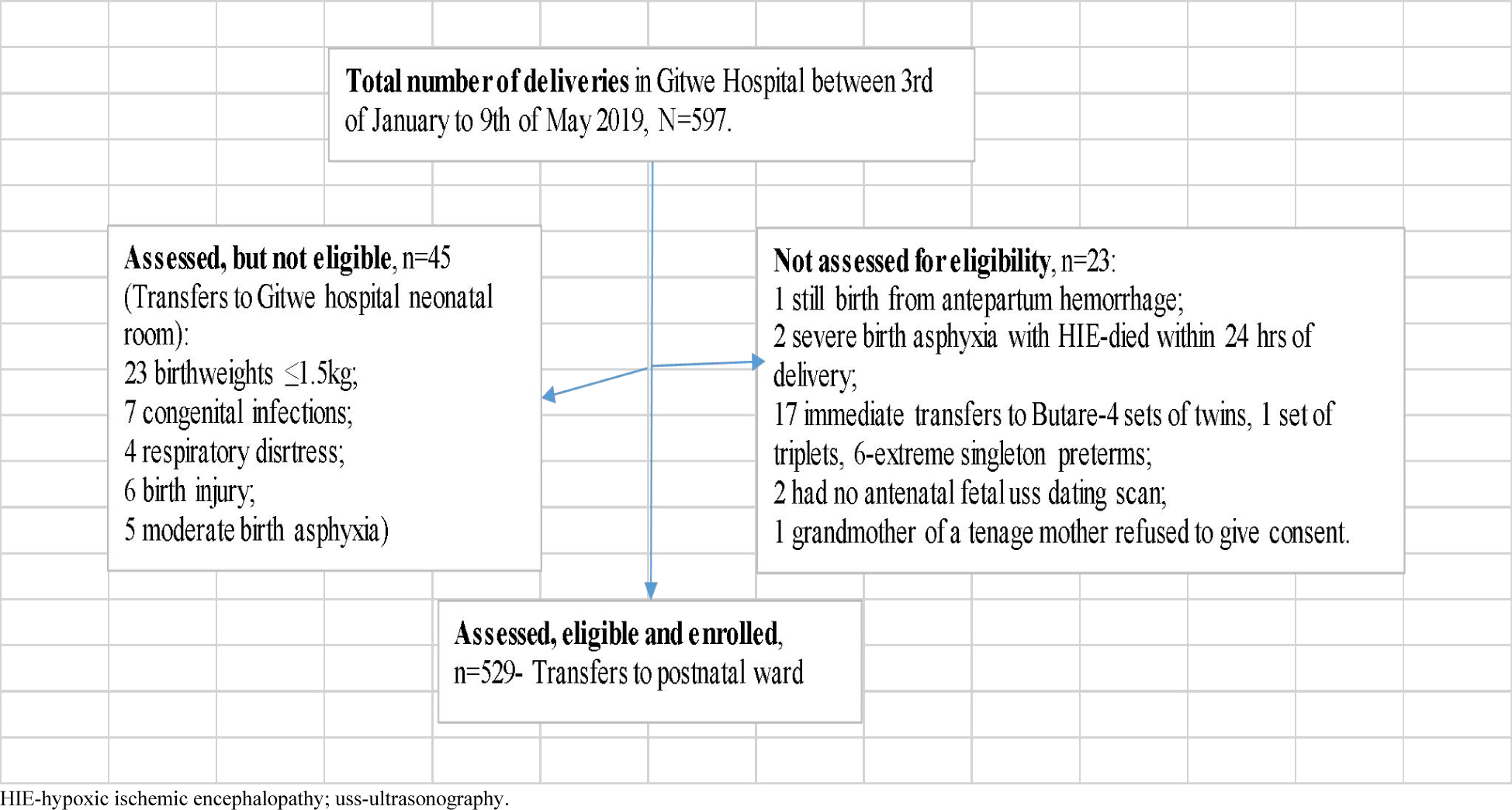
Flow of participants from admission to recruitment into study. At 6 weeks post delivery, 10 male infants (4 were born-LGA and 6 were born-AGA) were absent at the postnatal clinic and a further 7 babies (4 girls [1 was born-LGA, 1 born-AGA and 2 were born-SGA] and 3 boys [all were born-AGA]) were excluded from the analysis because of incomplete data. Hence, growth and feeding data were collected from a total of 512 babies at 6 weeks post-delivery. There were 242 girls (47.3%) and 270 boys (52.7%) at the 6 weeks postnatal clinic.

### Characteristics of babies associated with growth faltering at 6 weeks post-delivery

Overall, 16 (3.1%) out of the 512 babies were growth faltering according to static assessment in Table 1. The percentages in Table 1, shows that the girls were less likely (OR=0.36; 95% CI 0.10, 1.36) to falter in growth at 6 weeks post-delivery, compared to the boys, continuity correction p=0.204.

**Table 1:**
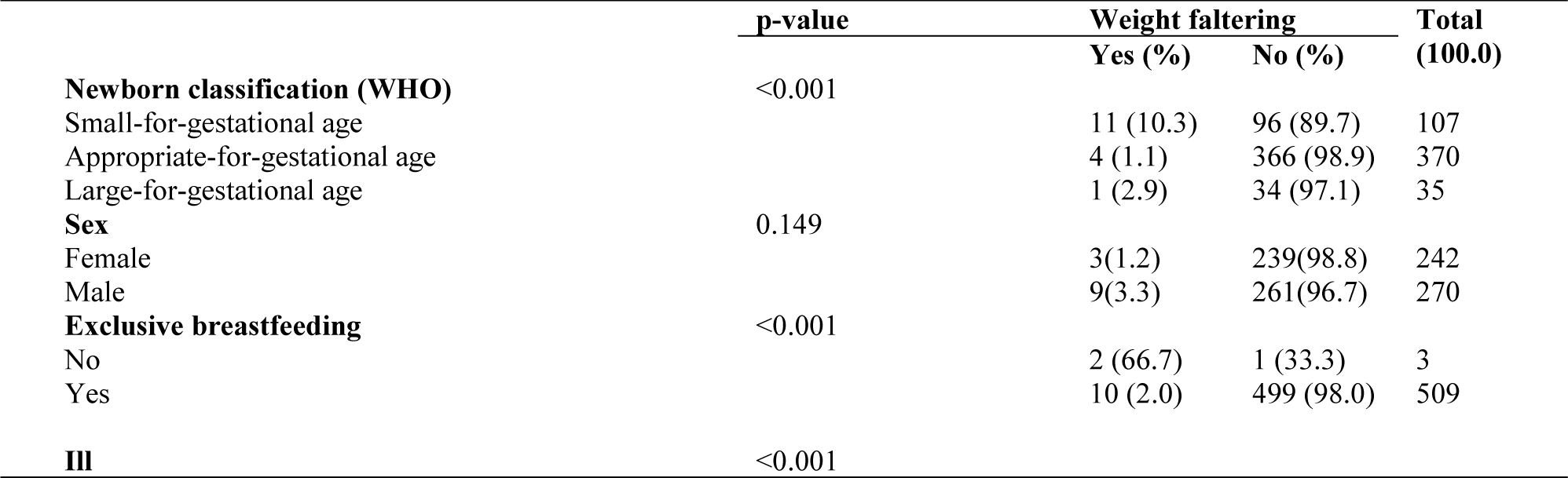
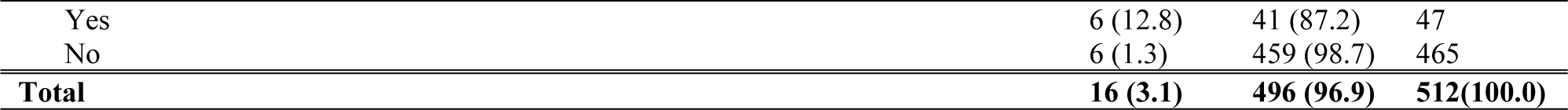
Crosstabulation of infant characteristics and weight faltering, n=512.

Three (0.6%) out of the 512 babies at 6weeks post-delivery were not exclusively breastfed. Two of these 3 babies, who were not on EBF were also offered water and the third one was cup-fed with whole cow milk. The baby who received cow milk, was also offered mashed potato porridge. The babies who were not exclusively breastfed were more likely to falter in growth (Fisher’s exact test was <0.001; OR=99.8; 95%CI was very wide).

107 SGA-born, 370 AGA-born and 35 LGA-born out of the 512 babies were analysed in the 6 weeks data. A higher percentage (10.3%) of the SGA babies, compared to 1.1% of AGA and 2.9% of the LGA faltered in weight, p<0.001. Of note, 7 (63.6%) out of the 11 SGA-born infants growth faltering were term asymmetrically grown SGA at birth.

However, according to incremental weight assessment, only 7 (1.4%) out of the 512 babies were weight faltering:SGA 2 (1.9%) out of 107; AGA 3 (0.8%) out of 370 and LGA 2 (5.7%) out of 35. This was the reverse of weight faltering pattern documented by static assessment.

47 (9.2%) of the 512 babies were reported ill in the first 6 weeks of life. Out of these 47 babies that were reported ill, 29 (5.7%) had respiratory illnesses; 7 (1.4%) had diarrhea illnesses, 4 (0.8%) were ill because of complications due to prematurity; 2 (0.4%) had sepsis and 1 (0.2%) each had respiratory illness and diarrhea, respiratory illness and malaria, jaundice, malaria and congenital heart disease. Those, who were reported ill were 11.20 (95% CI 3.45, 36.28) times more likely to falter in weight than those who had not fallen ill.

81 (15.8%) of the 512 babies were fed 8 times or less daily. These babies were 11.70 (95% CI 3.43, 39.85) times more likely to falter in growth, compared to those who were fed more frequently.

The median depression score of the 512 mothers was 7.0 (IQR 2,8). The mothers of babies who were faltering in weight had lower median depression score (0; IQR 0, 8.75) compared to those who were thriving (7.0; IQR 2.0, 8.0). However, the non-parametric independent t test statistics was not significant for the difference in the median depression scores between the 2 groups (p=0.716). There is likely to be a type II error, due to poor sample size of babies in the weight faltering group.

### Growth acceleration at 6 weeks post-delivery

When the infants were assessed for incremental growth, a higher percentage (81.3%) of the SGA babies, compared to the AGAs and LGAs were experiencing weight acceleration at 6 weeks post-delivery, p<0.001. Of note, the 78 (89.7%) out of the 87 SGA-born infants, growth accelerating at 6 weeks post-delivery, were symmetrically grown SGAs at birth. Only 6 (17.1%) of the LGA infants were growth accelerating, Table 2.

**Table 2:** Relationship between birth size and weight acceleration at 6 weeks post-delivery, n=512, p<0.001.

|  | Increment growth |  | Total |
| --- | --- | --- | --- |
|  | Not accelerating (%) | Accelerating (%) |  |
| SGA | 20 (18.7) | 87 (81.3) | 107 (100.0) |
| AGA | 121 (32.7) | 249 (67.3) | 370 (100.0) |
| LGA | 29 (82.9) | 6 (17.1) | 35 (100.0) |
| **Total** | **170 (33.2)** | **342 (66.8)** | **512 (100.0)** |

### Binary logistic regression analysis of factors associated with infant weight acceleration

The strongest factor was the birth size; the bigger the infant was at birth, the less likely the infant will weight accelerate at 6 weeks post-delivery; Wald 37.31, p<0.001. The LGA-born infant was the least likely to weight accelerate, compared to the SGA-born and AGA-born infants; OR 0.04; 95% CI 0.01, 0.10. High maternal depression score was associated with infant weight acceleration, Wald 14.37; p<0.001; OR 1.13; 95% CI 1.06, 1.21. Boys were less likely to weight accelerate compared to girls, p<0.001; Wald 12.94; OR 0.45; 95% CI 0.29, 0.70. Babies, who were adequately fed were more likely to weight accelerate compared to those who were poorly fed, Wald 11.62; p<0.001; OR 2.80; 95% CI 1.55, 5.05. Babies, who did not fall ill since birth were more likely to weight accelerate compared to those who had fallen ill, Wald 8.21; p=0.004; OR 2.73; 95% CI 1.37, 5.43.

**Table 3:**
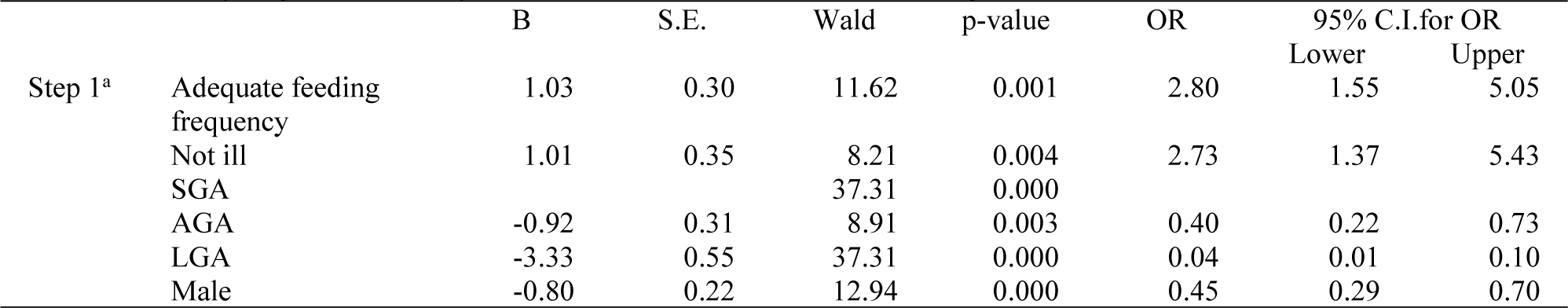
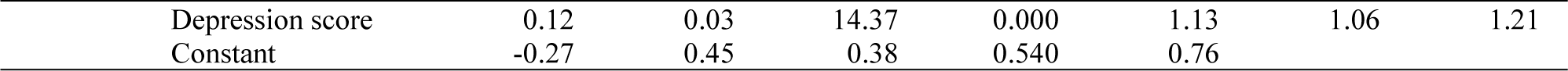
Binary regression analysis of infant characteristics and weight acceleration, n=512.

### Comparing static and incremental infant growth assessment

Conducting a Chi test using a 2 × 2 table, Fisher’s exact test was <0.001, suggesting a significant difference in the distribution of weight faltering in the findings of the 2 weight assessment methods (incremental vs static). McNemar test was significant at 0.022. Cohen Kappa measure of agreement between the 2 methods of weight assessment was moderate (0.424) and significant (p<0.001).

**Table 4:** Comparing incremental infant weight assessment to static infant growth assessment.

|  |  | Static infant weight assessment |  | Total |
| --- | --- | --- | --- | --- |
|  |  | Faltering (%) | Thriving (%) |  |
| Incremental infant weight assessment | Faltering | 5 (71.4) | 2 (28.6) | 7 |
|  | Thriving | 11 (2.2%) | 494 (97.8%) | 505 |
| Total |  | 16 (3.1) | 496 (96.9) | 512 |

## Discussion

This study estimated the incidence of weight faltering (based on both static and incremental weight assessment methods) among 512 infants, who were presented at the 6 weeks post-natal clinic. Inaddition, risk factors of postnatal weight faltering were identified and the 2 methods of weight assessment were compared. Of note, 10 infants out of the original 529 participants recruited from birth, were absent at the 6 weeks postnatal clinic.

The incidence of weight faltering at 6 weeks post-delivery by static assessment was higher than for incremental/interval assessment (3.1% versus 1.4%). The pattern of weight faltering also differed with both assessment methods. On the static assessment platform, the SGA-born infants were more inclined to weight faltering, while on the incremental assessment method, the SGA infants were found to be weight accelerating the most, compared to the AGA and LGA infants. The least accelerating group of infants in this study were the LGAs. This finding was in line with the principles of conditional weight gain, whereby the bigger babies tend to grow more slowly than the smaller sized babies from birth.^4^ Therefore, using only the static assessment method, could lead to increased tendency of SGA infants being mislabelled as weight faltering, aggressive feeding and subsequently metabolic problems later in life. The pathological weight faltering LGA infant is also likely to be missed.^4^

A small percentage (0.6%) of infants in this study were not exclusively breastfed. As such infants who were not adequately fed, not on exclusive breastfeeding, boys and those who fell ill were at increased risk of weight faltering. The birth size was also a significant determinant of infant growth at 6 weeks of life.^6^

Pellowski et al. observed a progressive decline in maternal median depressive symptom score from 9 [IQR: 6, 12] during pregnancy, to 8 [IQR: 5, 11] at both 10 weeks and 6 months postpartum, to 7 [IQR: 3.5, 11] at 12 months postpartum and then to 6 [IQR: 0,8] at 18 months postpartum, in South Africa. Though, the maternal depression scores in this study were lower compared to the South African study, the decline in maternal median depression symptom score from birth [7.0 (IQR 3,9)] to 6 weeks post-partum [7.0 (IQR 2,8)] was not as pronounced.^13^There is a strong link between stressful life events (which may include difficulties with lactation and newborn care) and maternal depression. It appears these mothers were still struggling with these challenges at 6 weeks post-delivery from birth.

One of the limitations in this study was the loss to follow up of infants at the MCH clinic.

### Conclusion and Recommendations

This study emphasizes the importance of assessing infant growth using both static and incremental measures to avoid mislabelling or missing infants with growth faltering. Health workers need to be trained to carry out incremental growth assessment in infants. Lactational and mental support programs should be strengthened in the rural MCH systems, to assist mothers to achieve pleasant experiences with breastfeeding and newborn care. Exclusive breastfeeding should be encouraged and this will in turn reduce the incidence of infant ill health. Home visits should be carried out for infants lost to follow up.

## Data Availability

All data produced in the present study are available upon reasonable request to the authors

## Author Contributions

The corresponding author (Dr Adenike Oluwakemi Ogah) conceived and designed the study, collected data and conducted data analysis, interpreted the results, and drafted the manuscript.

## Acknowledgements

The authors are extremely grateful to the participants involved in this study, to the staff of Gitwe Hospital and clinic in Rwanda and to the research team.

## Funding

This research was self-funded.

## Conflicts of Interest

The authors declare no conflict of interest.

## Notes

### Competing Interest Statement

The authors have declared no competing interest.

### Funding Statement

This study did not receive any funding

### Author Declarations

The Health Sciences Research Ethics Committee of the University of the Free State in South Africa gave the ethical approval for the collection of the primary data for the original study: Growth and growth faltering in a birth cohort in rural Rwanda: a longitudinal study with Ethical Clearance Number: UFS-HSD2018/1493/2901.

